# PathEQA: Feature-Graph-Guided Random Forests for Multianalyte External Quality Assessment

**DOI:** 10.64898/2026.08.20.26360960

**Authors:** Qing Li, Keying Yu

**Affiliations:** Department of Reference Measurement Laboratory, Shanghai Center for Clinical Laboratory, Shanghai, China

**Author notes:** Corresponding author: Qing Li.

**Keywords:** PathEQA, random forest, feature graph, graph-guided learning, external quality assessment, multianalyte quality assessment, catecholamine case study

## Abstract

External quality assessment (EQA) of multianalyte assays is commonly interpreted analyte by analyte, although many panels contain known relations among measured features that may reveal joint quality patterns. We propose PathEQA, a feature-graph-guided random forest framework in which a user-supplied graph can represent biochemical pathways, molecular interactions, shared measurement processes, or other domain relations. The same graph is allowed to influence feature representation, node-level candidate generation, and split selection, with an optional local grouped decision. We evaluated the framework in graph-aligned and graph-misspecified simulations and used a six-analyte catecholamine-related liquid chromatography-tandem mass spectrometry EQA data set as an illustrative case study (929 records from 58 laboratories and 117 complete multianalyte panels). In graph-aligned simulations, the grouped variant reduced test root mean squared error by 7.4-9.4% relative to ordinary random forest across training sizes of 60-240, whereas graph misspecification could worsen prediction. In the catecholamine case study, full PathEQA was comparable with ordinary random forest in laboratory-grouped cross-validation (RMSE 0.570 versus 0.569) and modestly better in the final-round temporal holdout (0.307 versus 0.318); a simpler static network-sampling baseline performed best. Dopamine-norepinephrine was the strongest pair, whereas dopamine-norepinephrine-epinephrine best estimated multianalyte failure burden. These results support a general conclusion: feature-graph guidance can improve small-sample multivariate quality assessment when the supplied structure is outcome-relevant, but graph relevance must be tested rather than assumed. Catecholamines serve here as a worked example rather than a restriction of the framework.

## 1 Introduction

Random forests are general-purpose ensemble learners that combine bootstrap aggregation with randomized feature selection at each node (Breiman, 2001). Their default construction deliberately treats candidate predictors as exchangeable. This design is robust and broadly applicable, but it does not exploit situations in which relations among predictors are known before training. Scientific data frequently provide such information as metabolic pathways, molecular interaction networks, engineering flow diagrams, spatial neighbourhoods, or measurement-process graphs. The central methodological question is not merely whether prior knowledge can be added to a forest, but where it should enter the learning algorithm and how its usefulness can be falsified.

Existing knowledge-guided forest approaches have mainly changed feature-sampling probabilities or used network information for pathway selection. Network-guided sampling can improve the recovery of connected modules, but it may leave prediction unchanged and can produce spurious selections when the supplied network is unrelated to the outcome (Hu & Szymczak, 2024). Knowledge-slanted forests similarly use network-derived probabilities to prioritize features, with benefits depending on the correctness of the prior information (Cantor et al., 2024). Pathway-hunting procedures use forests to rank predefined groups while retaining a largely conventional tree-growing mechanism (Chen & Ishwaran, 2013). These studies motivate a stronger design in which the same graph influences the representation, randomization, and optimization stages of tree learning, accompanied by explicit misspecification controls.

We therefore propose PathEQA, a feature-graph-guided random forest with a three-stage design. A fixed graph-informed representation provides neighbourhood context before tree growth; graph-conditioned candidate generation makes connected variables more likely to be considered together; and a structure-aware split objective can favour graph-consistent sequences of decisions. An optional grouped decision evaluates a local graph neighbourhood as a single decision unit. All components share the same user-supplied feature graph, and disabling graph guidance recovers the corresponding ordinary random forest. The framework is not tied to a specific analyte class: the graph can encode a metabolic pathway, a molecular interaction network, a measurement-process graph, or another scientifically motivated relation among predictors.

PathEQA is evaluated in controlled simulations and in a real external quality assessment (EQA) case study. EQA organizers usually assess each analyte separately against a consensus target. For multianalyte assays, this can miss joint quality patterns such as branch-wide bias, discordance between related measurements, or a laboratory-wide shift. Catecholamine-related LC-MS/MS EQA is used here as one concrete instantiation because the available data contain six connected analytes: dopamine (DA), norepinephrine (NE), epinephrine (E), 3-methoxytyramine (3-MT), normetanephrine (NMN), and metanephrine (MN). The methodological framework itself is intended for any multianalyte setting in which a defensible feature graph can be supplied.

The paper makes four contributions:

- PathEQA, a random-forest construction that uses one feature graph at the representation, candidate-generation, and split-selection levels, with an optional local grouped-decision extension.
- High-level degeneration, graph-label equivariance, and computational properties that clarify what the structural prior can and cannot guarantee without disclosing implementation-enabling mathematics.
- A falsification-oriented experimental design including ablation, static-sampling, shuffled-graph, and graph-misspecified controls rather than assuming that a supplied domain graph is predictive.
- A catecholamine EQA case study that identifies useful analyte combinations and laboratory-level quality phenotypes while illustrating a broader point: a biological graph and a measurement-error graph need not be identical.

## 2 Related work

Breiman’s random forest uses bootstrap samples, randomized node-level candidate features, and aggregation to reduce variance while retaining nonlinear interactions (Breiman, 2001). Subsequent theoretical work established consistency under particular model assumptions and clarified the interaction between sparsity, randomization, and tree structure (Scornet et al., 2015). The present work preserves bootstrap aggregation and recursive partitioning but changes the feature representation, candidate distribution, and split objective.

Graph-based prior knowledge has been incorporated into learning through regularization, spectral transforms, group penalties, and biased feature selection. Graph-Laplacian methods exploit the positive semidefinite operator L = D − A to impose smoothness over related variables or samples (Belkin et al., 2006). In forests, prior networks have commonly been summarized as static feature weights. Hu and Szymczak (2024) showed that this strategy can improve connected-module recovery but not necessarily prediction, and can emphasize hub variables spuriously. Cantor et al. (2024) used random-walk-derived feature probabilities in small-sample genomics. Graph-guided and pathway-hunting forests instead focus on identifying connected feature sets or ranking pathways (Chen & Ishwaran, 2013; Pfeifer et al., 2021).

PathEQA differs in where and how domain structure enters the forest. The feature graph is not used only as a preprocessing filter or a single static probability vector. Instead, one graph informs the input representation, the node-level randomized search, and the local split preference. This coupling also creates a clear ablation design: representation only; representation plus graph-conditioned sampling; full PathEQA; and the optional grouped extension. In this priority-establishing preprint, the conceptual architecture and empirical behaviour are disclosed while implementation-enabling mathematical details are intentionally omitted.

## 3 PathEQA: feature-graph-guided random forest

### 3.1 Problem definition and graph notation

Let the predictor matrix contain n observations and p features, with a regression response and a user-supplied undirected feature graph linking predictors that are considered related by prior knowledge. The graph may be binary or weighted and may represent biochemical adjacency, shared analytical steps, molecular interactions, or another domain structure. PathEQA uses the same graph consistently across its graph-dependent components.

For the six-analyte catecholamine case study, the nodes are DA, NE, E, 3-MT, NMN, and MN, with edges DA-NE, NE-E, DA-3-MT, NE-NMN, and E-MN. These edges encode immediate biochemical relations among the measured analytes. This six-node pathway is one application graph; other applications can substitute a different feature graph without changing the overall learning framework.

### 3.2 Graph-informed representation

#### Disclosure note for Section 3

This public preprint is intended to establish the conceptual architecture, scope, and empirical findings of PathEQA while preserving implementation-specific intellectual property. It therefore omits the exact transformation operator, node-level sampling kernel, split-score formula, grouped aggregation function, pseudocode, and exact graph-guidance tuning constants. These details would be required for straightforward reimplementation and are reserved for a later technical release.

Before tree growth, the predictors are passed through a fixed graph-informed representation that blends each feature with information from its local graph context. The transformation is determined by the supplied graph rather than estimated from the response, and it is chosen to be numerically stable while preserving graph-constant components. The exact operator and parameterization are withheld in this preprint version. In the EQA case study, a dimensionless log-ratio transformation is applied before the graph-informed representation so that analytes with different physical units and concentration scales can be considered jointly.

### 3.3 Graph-conditioned candidate sampling

At each tree node, PathEQA modifies the ordinary random-forest candidate search so that the candidate set is conditioned on a randomly selected location in the feature graph. Connected predictors are therefore more likely to be considered together than under purely uniform feature sampling, while stochastic exploration and a graph-free fallback are retained. The exact sampling kernel, probability schedule, and budget-handling rules are intentionally withheld in this preprint version.

### 3.4 Structure-aware split scoring

For each candidate split, PathEQA evaluates the ordinary predictive improvement together with a bounded structural preference derived from the feature graph and the sequence of features already used on the current root-to-node path. This makes the structural preference conditional on the evolving tree rather than a fixed marginal feature weight. The precise normalization, graph-history function, and weighting rule are withheld in this preprint version.

### 3.5 Optional local grouped decision

The optional grouped extension changes the decision unit from a single graph-informed feature to a compact local summary of that feature and its immediate graph neighbourhood. This expands the split search space and is therefore reported separately from the core model. The exact aggregation function is intentionally withheld in this preprint version.

### 3.6 Degeneration, equivariance, and complexity

#### Degeneration

When all graph-dependent components are disabled, PathEQA reduces exactly to the corresponding ordinary random-forest implementation. This keeps the graph-free baseline algorithmically aligned with the proposed model rather than relying on a separately tuned comparator.

#### Graph-label equivariance

Simultaneously relabeling the predictor columns and the supplied graph relabels all graph-dependent operations in the same way; after mapping feature labels back, the fitted-forest distribution is unchanged. The method therefore depends on graph structure rather than arbitrary column order.

#### Computational behaviour

The added operations are local to the feature graph and are small relative to threshold evaluation during tree growth for the low-dimensional settings studied here. Sparse graph representations can be used for larger feature sets. Exact implementation-level complexity constants are not disclosed in this version.

**Fig. 1.**
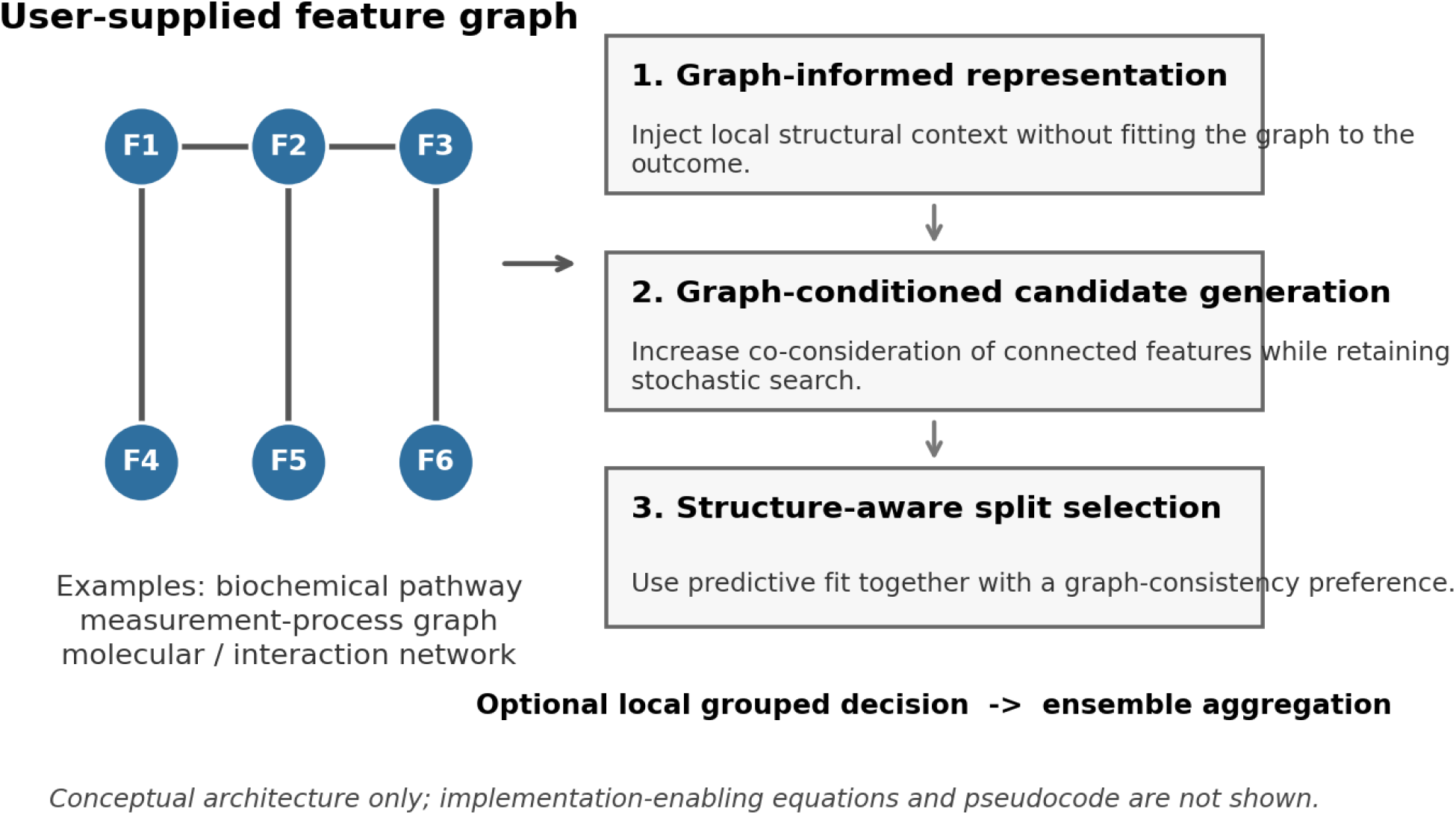
Conceptual overview of PathEQA. A user-supplied feature graph can encode biochemical, molecular, analytical-process, or other domain relations. The same graph informs three conceptually distinct stages of the forest: feature representation, candidate generation, and split selection. An optional local grouped decision can also be enabled. The catecholamine graph used later is one application instance. Implementation-enabling equations and pseudocode are intentionally not shown in this preprint version.

## 4 Experimental design

### 4.1 Synthetic experiments

We constructed six-dimensional regression problems using a graph with the same size and connectivity pattern as the case-study graph but with generic feature labels. Predictors were generated with a graph-aligned correlation structure. In the aligned setting, the response contained nonlinear main and interaction effects concentrated on graph-connected features; in the graph-misspecified setting, the outcome signal was reassigned to nonedges while the algorithm continued to receive the original graph. The exact covariance construction and response-generating functions are withheld in this preprint version. Training sizes were n = 60, 120, and 240; each replicate used an independent test set of 1,000 observations, and six replicates were run for each setting.

We compared ordinary random forest, representation only, representation plus graph-conditioned sampling, full PathEQA, PathEQA with grouped decisions, static network sampling, and a label-permuted graph. All model settings were prespecified and held fixed within each experiment family; exact graph-guidance constants and implementation-specific tree settings are intentionally withheld in this preprint version. Primary metrics were test RMSE and R^2^. Graph coherence was defined as the proportion of consecutive split-feature pairs connected by the supplied graph, excluding pairs in which a node or its parent was a leaf.

### 4.2 Catecholamine EQA case-study data

The EQA data contained 929 laboratory–analyte–round records from 58 laboratories across six rounds between May 2023 and September 2025. Each record contained results for five EQA materials. The measured analytes were 3-MT, DA, E, MN, NE, and NMN. Only 3-MT, MN, and NMN were present in the two 2023 rounds; all six analytes were available from 2024 onward. Complete six-analyte laboratory–round panels were therefore used for the graph analysis, yielding 117 panels: 22 in May 2024, 21 in September 2024, 37 in May 2025, and 37 in September 2025.

For each analyte a, round r, and material s, the all-participant median M(a,r,s) was the consensus target. A dimensionless input was calculated as x = log[reported result/M(a,r,s)]. A material result was acceptable when the untransformed ratio was between 0.75 and 1.25. An analyte was classified as passing when at least four of five material results were acceptable. The panel-level regression target was failed-analyte burden, the number of failed analytes from 0 to 6. This target was chosen because it represents multianalyte quality burden rather than repeating six independent binary grades.

Each laboratory–round panel was treated as a bag of five material-level six-analyte vectors. A forest generated one burden prediction per material, and the five predictions were averaged to obtain the panel estimate. This preserves the algorithm’s six-node input while using all five EQA materials. All instances from a laboratory remained in the same cross-validation partition.

### 4.3 Baselines, validation, and metrics

The EQA comparison used the same algorithmic variants as the simulation. Static network sampling used fixed graph-derived candidate weights but did not use the node-conditioned neighbourhood mechanism or the structure-aware split preference. The shuffled-graph control permuted analyte labels on the adjacency structure. Hyperparameters were fixed before the final temporal evaluation; the exact implementation-specific values are intentionally withheld in this preprint version.

The primary validation was five-fold cross-validation grouped by laboratory code, repeated over three forest seeds. Performance was aggregated at the laboratory–round panel level. A temporal holdout trained on complete panels from May 2024 through May 2025 and tested on September 2025. Metrics were RMSE and R^2^ for burden estimation and ROC area under the curve (AUC) and average precision (AP) for detecting any panel with at least one failed analyte. Results are reported as mean ± standard deviation across seeds.

### 4.4 Indicator combinations and failure phenotypes

To determine which analytes were jointly informative, feature importance was accumulated from normalized split gains in the full model over ten independent seeds. Candidate two- and three-analyte subsets were then refitted and evaluated by laboratory-grouped cross-validation over five seeds. Pair and triple results were interpreted as predictive combinations, not causal biochemical effects.

Failure phenotypes were explored among the 37 complete panels with at least one failed analyte. For each analyte we calculated the median signed log-ratio across five materials and the root-mean-square log-ratio, producing 12 panel features. K-means clustering with k = 3 was selected as the smallest solution yielding technically distinct patterns. Separately, edge discordance was the mean absolute difference in log-ratios across the five graph edges and five EQA materials. A robust high-discordance threshold was the median plus three scaled median absolute deviations among panels in which all six analytes passed.

### 4.5 Software

Analyses were implemented in Python using NumPy, pandas, SciPy, scikit-learn utilities for metrics and validation, and a custom tree/forest implementation for the graph-guided components of PathEQA. Matplotlib was used for figures. The custom source code and implementation-enabling specification are not included in this public preprint version.

**Table 1.** Algorithmic variants and graph-guidance components enabled in each model.

| Variant | Graph-informed representation | Graph-conditioned sampling | Structure-aware split score | Grouped decision | Purpose |
| --- | --- | --- | --- | --- | --- |
| Ordinary RF | No | No | No | No | Exact baseline |
| Projection only | Yes | No | No | No | Representation contribution |
| Projection + sampling | Yes | Yes | No | No | Conditional randomization contribution |
| Full PathEQA | Yes | Yes | Yes | No | Three-stage core model |
| PathEQA + grouped split | Yes | Yes | Yes | Yes | Expanded decision unit |
| Static network sampling | No | Static weights | No | No | Closest sampling-only baseline |
| Shuffled graph | Yes | Yes | Yes | No | Graph-specificity control |

## 5 Results

### 5.1 Graph-aligned and graph-misspecified simulations

In graph-aligned regression, all graph-representation variants improved RMSE relative to ordinary random forest, and the grouped-split extension was best at every training size (Fig. 2; Table 2). At n = 60, RMSE decreased from 1.198 ± 0.061 for ordinary random forest to 1.085 ± 0.046 for the grouped model, a 9.4% reduction. Corresponding reductions were 7.4% at n = 120 and 8.7% at n = 240. Full PathEQA without grouped splits increased graph coherence from 0.350 to 0.406 at n = 60 and from 0.363 to 0.403 at n = 240. Thus the structure-aware score changed the learned split topology even when its predictive gain over representation alone was modest.

**Table 2.** Graph-aligned simulation performance.

| n | Model | RMSE | R<sup>2</sup> | Graph coherence |
| --- | --- | --- | --- | --- |
| 60 | Ordinary RF | 1.198 ± 0.061 | 0.512 ± 0.048 | 0.350 ± 0.031 |
| 60 | Projection only | 1.131 ± 0.046 | 0.566 ± 0.034 | 0.348 ± 0.037 |
| 60 | Projection + sampling | 1.129 ± 0.050 | 0.567 ± 0.040 | 0.375 ± 0.030 |
| 60 | Full PathEQA | 1.130 ± 0.049 | 0.566 ± 0.040 | 0.406 ± 0.036 |
| 60 | PathEQA + grouped split | 1.085 ± 0.046 | 0.600 ± 0.037 | 0.421 ± 0.030 |
| 120 | Ordinary RF | 1.120 ± 0.033 | 0.567 ± 0.022 | 0.354 ± 0.016 |
| 120 | Projection only | 1.068 ± 0.029 | 0.606 ± 0.018 | 0.350 ± 0.028 |
| 120 | Projection + sampling | 1.085 ± 0.030 | 0.593 ± 0.022 | 0.386 ± 0.028 |
| 120 | Full PathEQA | 1.083 ± 0.030 | 0.595 ± 0.021 | 0.402 ± 0.031 |
| 120 | PathEQA + grouped split | 1.036 ± 0.050 | 0.629 ± 0.023 | 0.405 ± 0.015 |
| 240 | Ordinary RF | 1.081 ± 0.036 | 0.600 ± 0.013 | 0.363 ± 0.019 |
| 240 | Projection only | 1.042 ± 0.029 | 0.629 ± 0.007 | 0.346 ± 0.025 |
| 240 | Projection + sampling | 1.042 ± 0.028 | 0.629 ± 0.012 | 0.380 ± 0.021 |
| 240 | Full PathEQA | 1.044 ± 0.026 | 0.627 ± 0.010 | 0.403 ± 0.018 |
| 240 | PathEQA + grouped split | 0.987 ± 0.022 | 0.666 ± 0.014 | 0.383 ± 0.017 |

**Fig. 2.**
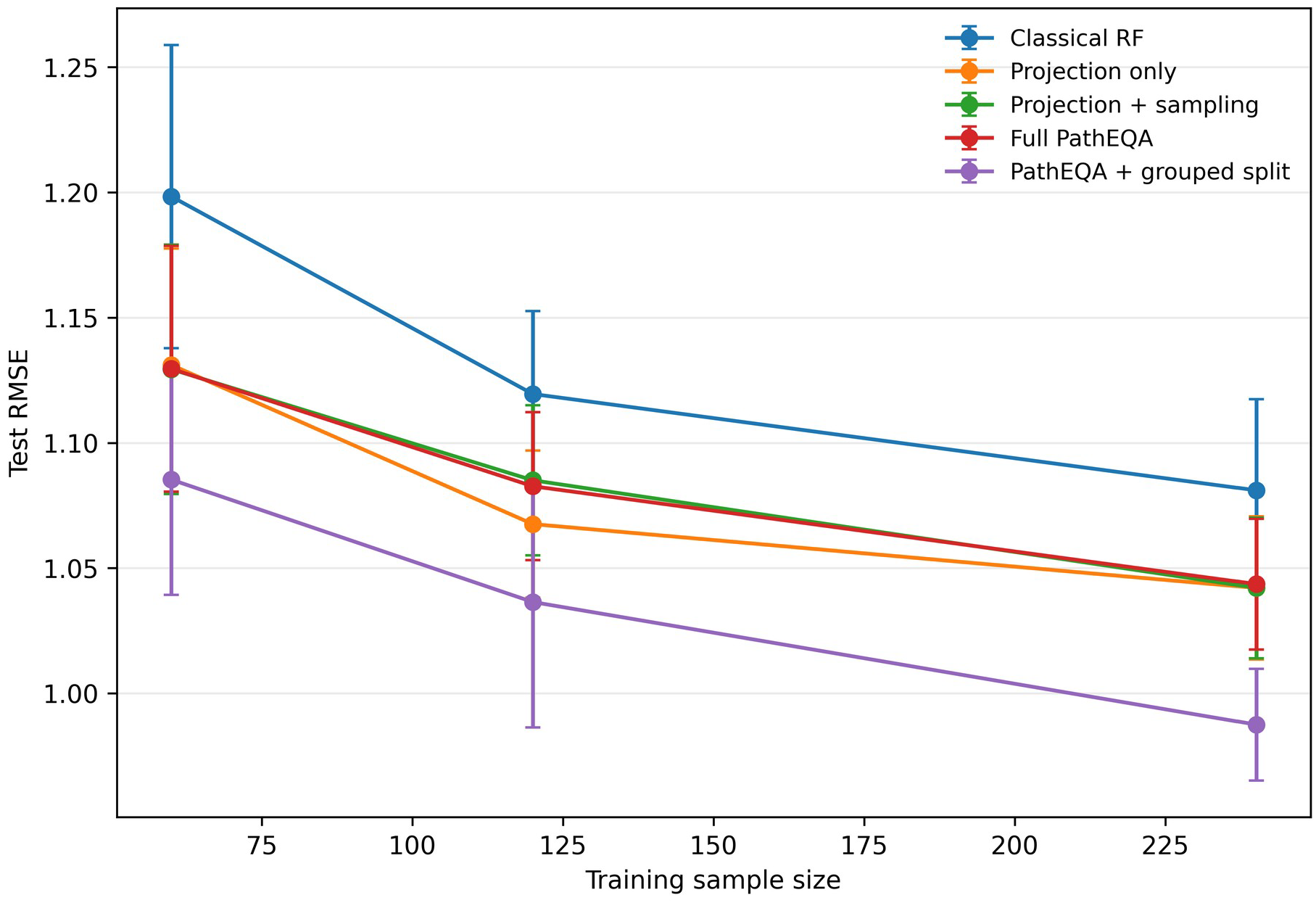
Test RMSE in graph-aligned synthetic regression. Points are means over six replicates and error bars are standard deviations

Graph guidance did not provide a universal advantage. In the graph-misspecified experiment at n = 120, ordinary random forest achieved RMSE 1.117 ± 0.036, whereas full PathEQA yielded 1.142 ± 0.040 and the grouped extension 1.171 ± 0.029. These results confirm that a mathematically well-behaved graph-informed representation does not imply predictive robustness to an incorrect feature graph. A graph-guided method therefore requires a negative control and should not be evaluated only in aligned simulations.

### 5.2 EQA data structure and failure burden

Among all 929 records, analyte counts were 194 for MN, 194 for NMN, 175 for 3-MT, 123 for E, 122 for NE, and 121 for DA. Of 117 complete six-analyte panels, 37 contained at least one failed analyte. DA accounted for 31 failures, followed by E (11), NE (6), 3-MT (4), MN (2), and NMN (1). Because several analytes could fail in the same panel, these counts exceed the number of failed panels.

### 5.3 Predictive performance and graph coherence

Full PathEQA did not improve the primary laboratory-grouped cross-validation RMSE over ordinary random forest: 0.570 ± 0.007 versus 0.569 ± 0.005 (Fig. 3a; Table 3). The grouped-split extension was slightly better for burden estimation (0.567 ± 0.005) but reduced any-failure discrimination. The static network-sampling baseline had the lowest cross-validated RMSE, 0.553 ± 0.003, with R^2^ 0.616 ± 0.004. AUC values were high for all core variants, ranging from 0.951 to 0.955, indicating that much of the any-failure signal was already available without graph guidance.

**Table 3.** EQA predictive performance.

| Model | Grouped-CV RMSE | Grouped-CV $R^2$ | Grouped-CV AUC | 2025.9 RMSE | 2025.9 AUC |
| --- | --- | --- | --- | --- | --- |
| Ordinary RF | $0.570 \pm 0.005$ | $0.593 \pm 0.008$ | $0.953 \pm 0.003$ | $0.317 \pm 0.006$ | $0.963 \pm 0.005$ |
| Projection only | $0.571 \pm 0.003$ | $0.590 \pm 0.004$ | $0.954 \pm 0.004$ | $0.312 \pm 0.008$ | $0.975 \pm 0.002$ |
| Projection + pathway sampling | $0.570 \pm 0.007$ | $0.592 \pm 0.010$ | $0.952 \pm 0.002$ | $0.309 \pm 0.006$ | $0.962 \pm 0.002$ |
| Full PathEQA | $0.570 \pm 0.007$ | $0.592 \pm 0.009$ | $0.952 \pm 0.001$ | $0.307 \pm 0.006$ | $0.962 \pm 0.002$ |
| PathEQA + grouped split | $0.567 \pm 0.005$ | $0.596 \pm 0.007$ | $0.934 \pm 0.007$ | $0.322 \pm 0.012$ | $0.952 \pm 0.008$ |
| Static network sampling | $0.553 \pm 0.003$ | $0.616 \pm 0.003$ | $0.951 \pm 0.003$ | $0.299 \pm 0.007$ | $0.967 \pm 0.005$ |
| Shuffled graph | $0.570 \pm 0.011$ | $0.591 \pm 0.015$ | $0.955 \pm 0.001$ | $0.300 \pm 0.003$ | $0.980 \pm 0.004$ |

**Table 4.** Repeated laboratory-grouped cross-validation of selected analyte combinations.

| Combination | RMSE | R<sup>2</sup> | AUC | Average precision | Primary interpretation |
| --- | --- | --- | --- | --- | --- |
| DA+NE | $0.561 \pm 0.004$ | $0.605 \pm 0.005$ | $0.947 \pm 0.002$ | $0.939 \pm 0.001$ | Best pair |
| DA+E | $0.601 \pm 0.003$ | $0.546 \pm 0.005$ | $0.923 \pm 0.001$ | $0.934 \pm 0.002$ | Strong nonedge comparator |
| DA+3-MT | $0.629 \pm 0.004$ | $0.503 \pm 0.006$ | $0.948 \pm 0.003$ | $0.912 \pm 0.004$ | Direct metabolic pair |
| DA+NE+E | $0.549 \pm 0.002$ | $0.621 \pm 0.003$ | $0.946 \pm 0.002$ | $0.943 \pm 0.001$ | Best burden estimation |
| DA+NE+NMN | $0.566 \pm 0.003$ | $0.598 \pm 0.005$ | $0.961 \pm 0.002$ | $0.943 \pm 0.002$ | Best any-failure discrimination |
| DA+NE+3-MT | $0.578 \pm 0.004$ | $0.581 \pm 0.006$ | $0.950 \pm 0.001$ | $0.913 \pm 0.002$ | Catecholamine plus DA metabolite |

**Fig. 3.**
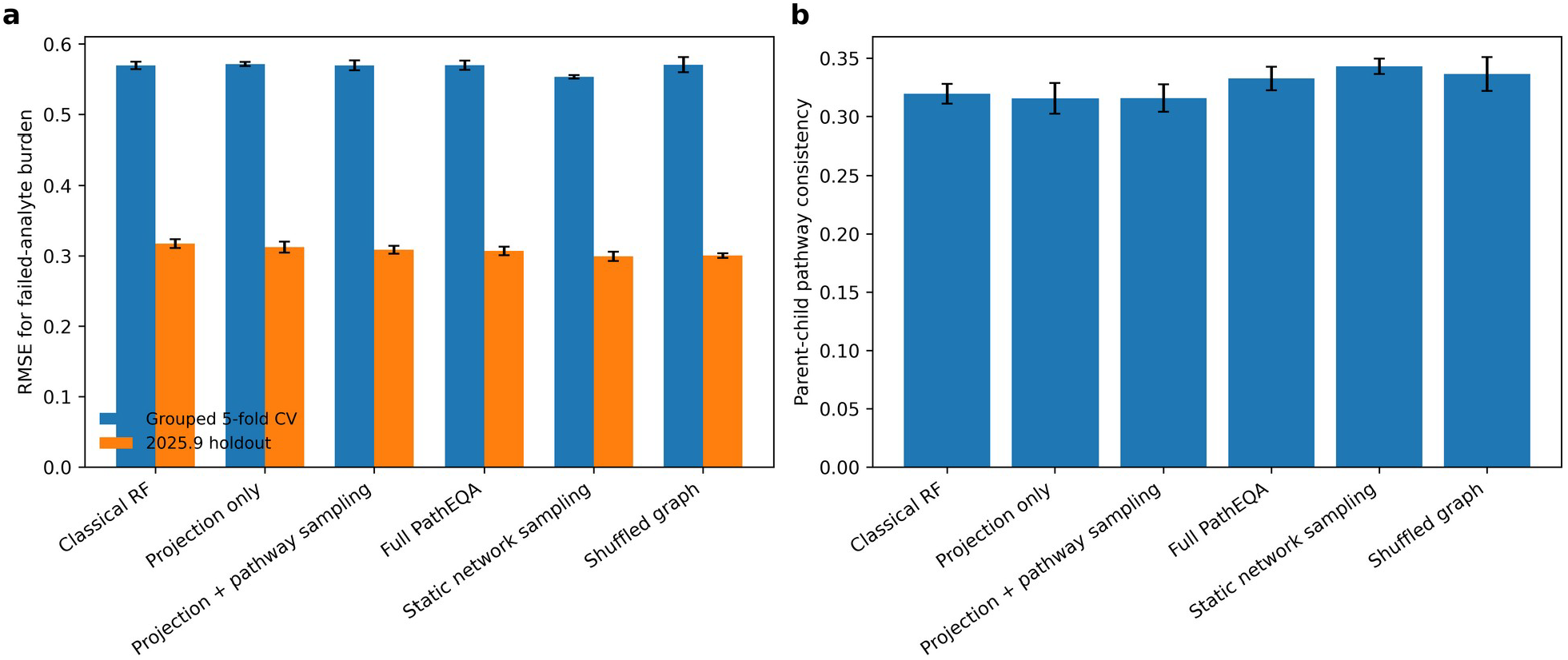
Performance of PathEQA and comparison models on the six-analyte EQA data. a RMSE in laboratory-grouped cross-validation and the September 2025 temporal holdout. b Parent–child graph coherence in fitted trees. Error bars are standard deviations over independent forest seeds

In the September 2025 temporal holdout, full PathEQA achieved RMSE 0.307 ± 0.006 compared with 0.318 ± 0.006 for ordinary random forest. Static network sampling again had the lowest RMSE (0.299 ± 0.007), and the shuffled graph was similar (0.301 ± 0.003). The fact that a permuted graph matched or exceeded the biochemical pathway graph indicates that the observed temporal gain cannot be attributed uniquely to the known metabolic edges.

Full PathEQA increased parent–child graph coherence in grouped cross-validation from 0.320 ± 0.008 to 0.333 ± 0.010 (Fig. 3b). The grouped-split extension reached 0.358 ± 0.003. Graph coherence therefore responded to the algorithmic prior, but higher coherence did not monotonically translate into lower prediction error.

### 5.4 Informative analytes and joint indicator sets

Normalized split-gain importance in full PathEQA was highest for DA (0.346 ± 0.021), followed by E (0.212 ± 0.014), NE (0.131 ± 0.011), 3-MT (0.122 ± 0.015), NMN (0.115 ± 0.016), and MN (0.074 ± 0.009) (Fig. 4a). The most frequent connected parent–child split pairs included DA–NE and DA–3-MT, but the nonedge DA–E pair was also frequent. This again suggests that biochemical adjacency captures part, but not all, of the analytical dependence structure.

**Fig. 4.**
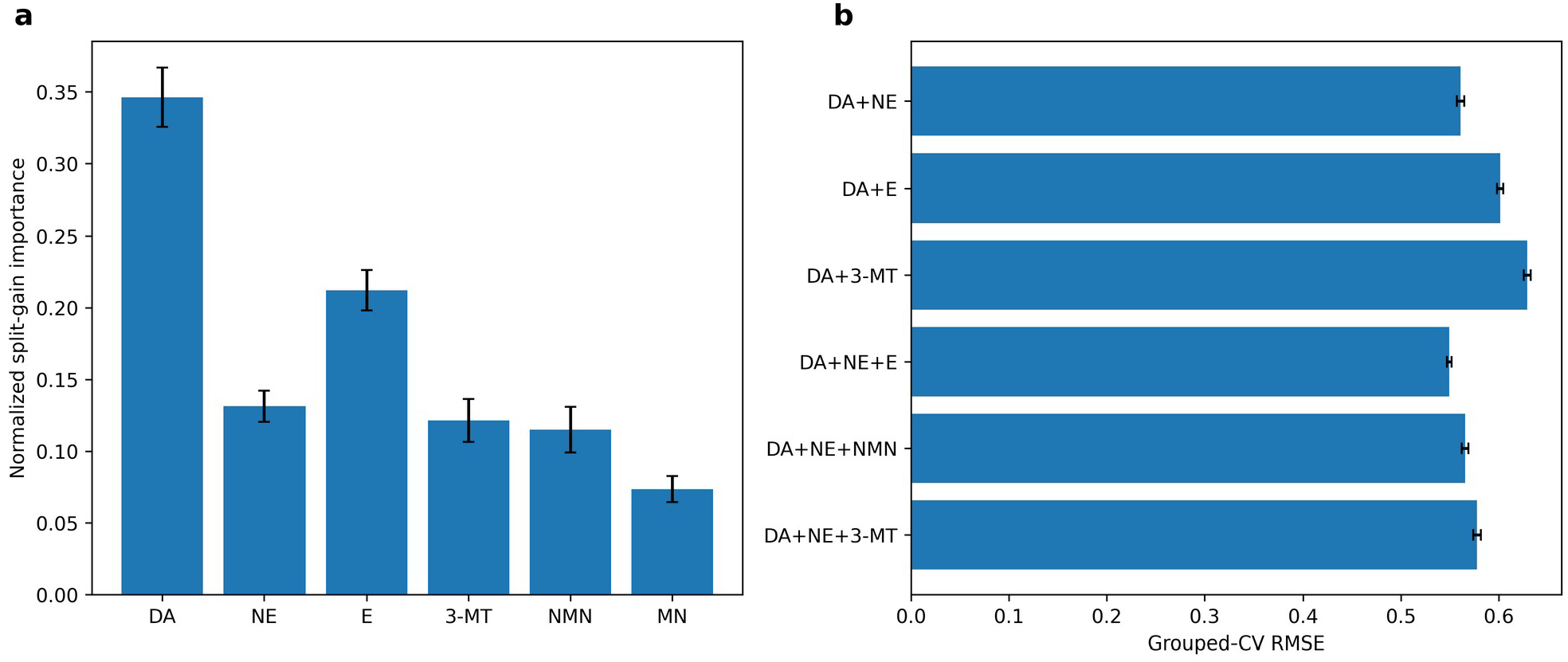
Analyte contributions and reduced indicator sets. a Normalized split-gain importance in full PathEQA. b Grouped-CV RMSE for selected two- and three-analyte subsets. Error bars are standard deviations over independent forest seeds

Among two-analyte models, DA–NE had the lowest grouped-CV RMSE (0.561 ± 0.004) and an AUC of 0.947 ± 0.002. For three analytes, DA–NE–E best estimated continuous failed-analyte burden (RMSE 0.549 ± 0.002; R^2^ 0.621 ± 0.003), whereas DA–NE–NMN provided the highest any-failure AUC (0.961 ± 0.002). The optimal combination therefore depended on the operational objective: estimating total burden versus screening for any quality failure.

### 5.5 Multianalyte quality phenotypes and graph-based warnings

Clustering the 37 failed complete panels produced three interpretable patterns (Fig. 5a). The largest phenotype (n = 28) was DA-dominant mixed or positive bias, with mean failed-analyte burden 1.25; 23 panels failed DA and seven failed E. A second phenotype (n = 8) showed catecholamine-branch low bias, with mean signed log-ratios of −0.525 for DA, −0.158 for NE, and −0.194 for E and mean burden 1.75. One severe global-distortion panel failed all six analytes and showed marked negative shifts across most nodes. These clusters describe result patterns and do not establish root causes.

**Fig. 5.**
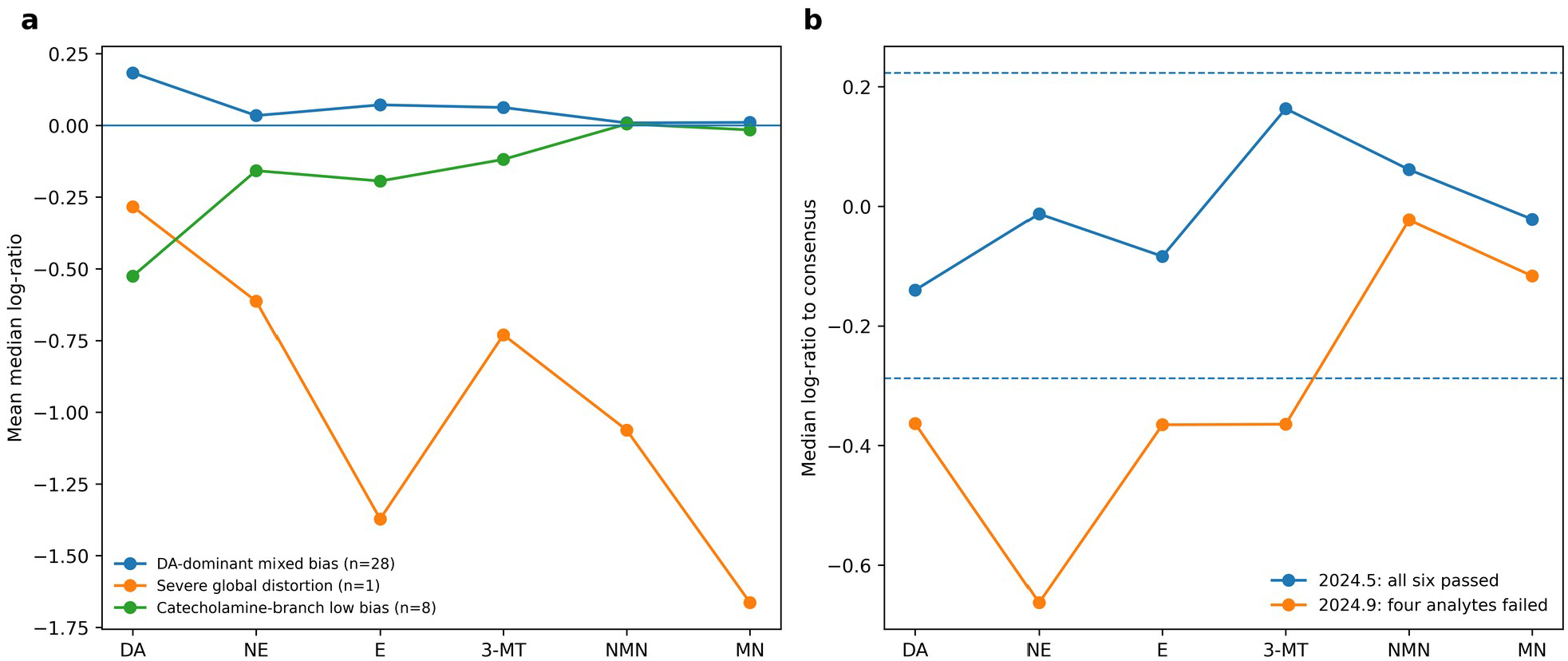
Multianalyte quality patterns. a Mean signed log-ratio profiles for three failure phenotypes. b An illustrative laboratory that passed all six analytes in one round despite high structural discordance and failed four analytes in the subsequent round. Dashed lines show log(0.75) and log(1.25)

Edge discordance was substantially larger in failed than all-pass panels: median 0.254 versus 0.085. The robust all-pass threshold was 0.175. Two panels passed all six analytes while exceeding this threshold. One had a subsequent round available: its initial panel passed all six analytes but displayed strong cross-edge disagreement; in the next round DA, NE, E, and 3-MT failed (Fig. 5b). This single case illustrates the type of hidden multianalyte warning that a graph-guided EQA report could surface, but the available number of rounds was insufficient to validate next-round forecasting.

## 6 Discussion

PathEQA was designed to answer a general machine-learning question with direct relevance to multianalyte quality assessment: how can one user-supplied feature graph influence a forest at more than one stage without hiding the consequences of an incorrect prior? The construction couples graph-informed representation, graph-conditioned randomized search, and structure-aware split optimization. The graph can be biological, analytical, or otherwise domain-defined; catecholamine EQA is used here as an illustrative case rather than as the scope boundary of the method. A graph-free degeneration keeps the baseline well defined, while shuffled-graph and graph-misspecified experiments separate structural preference from genuine predictive value.

The graph-aligned simulations show the intended regime of benefit. The graph-informed representation delivered most of the predictive improvement, while graph-conditioned sampling and structure-aware scoring increased graph coherence. Grouped decisions added a consistent RMSE reduction, suggesting that local graph summaries can be useful when the response depends on connected feature combinations and the sample size is small. This result is consistent with the broader idea that prior knowledge is most helpful when the data alone are insufficient to recover interactions. However, the misspecified simulation shows the corresponding cost: enforcing a wrong feature graph can increase error even when the representation itself is numerically stable.

The catecholamine EQA analysis provides a deliberately difficult real-world case study rather than a showcase data set. Full PathEQA was not superior to ordinary random forest in the primary grouped cross-validation, although it was modestly better in the temporal holdout and produced more graph-coherent split sequences. A simple static network-sampling baseline performed best for burden estimation, and a shuffled graph performed similarly or better in the temporal holdout. The correct interpretation is therefore not that pathway information is universally useful, but that the most relevant feature graph depends on the error-generating mechanism. In laboratory quality assessment, shared calibration, extraction, derivatization, internal standards, chromatographic interference, ionization, unit conversion, and data-processing steps can couple analytes that are not adjacent in metabolism. A measurement-process graph may therefore be more relevant to analytical quality than a pure metabolic pathway graph.

The indicator-combination analysis is operationally useful for an EQA organizer. DA–NE was the strongest two-analyte set and represents a direct pathway edge. The best burden-estimation triple, DA–NE–E, follows the catecholamine branch. By contrast, DA–NE–NMN best discriminated whether any failure was present. This difference matters: an organizer seeking to estimate the number of affected analytes may choose a different reduced panel from one seeking a sensitive screening flag. The high importance of DA is also consistent with the observed failure distribution, but the joint models show that DA should not be interpreted in isolation.

The failure phenotypes extend conventional single-analyte grading. A DA-dominant pattern may prompt analyte-specific review; branch-wide low bias may prompt review of shared extraction, calibration, or sensitivity; and severe global distortion may prompt unit, dilution, transcription, or system-wide investigations. These are triage hypotheses rather than automated root-cause diagnoses. Confirmatory information such as calibration curves, internal-standard responses, chromatograms, recovery experiments, and participant comments would be required before assigning a cause.

The study also illustrates a broader evaluation principle for knowledge-guided machine learning. Structural coherence is not a substitute for prediction, and predictive improvement alone does not prove that the supplied graph is correct. A convincing evaluation should report both, include graph-free and simpler graph baselines, and compare with random or deliberately incorrect graphs. Prior-knowledge methods should be viewed as conditional learners: their usefulness depends on alignment among the graph, the data-generating mechanism, and the prediction target. This conclusion agrees with prior evaluations showing that network-guided random forests can improve connected-module recovery while failing to improve prediction or inducing spurious selections under network misspecification (Hu & Szymczak, 2024).

Several limitations constrain the present conclusions. First, the real study contained only 117 complete panels and four rounds with all six analytes. The temporal holdout therefore represents one future round, not external validation. Second, the panel target was contemporaneous failed-analyte burden. The structural-warning case motivates, but does not validate, next-round prediction. Third, the consensus median and ±25% criterion define the outcome; alternative EQA specifications may change the failure distribution. Fourth, the supplied graph contained only six measured nodes and omitted unmeasured intermediates. Fifth, one label-permuted graph does not characterize the full random-graph null distribution. Sixth, hyperparameters were fixed rather than selected in a fully nested procedure. Finally, no independent EQA scheme was available to test transportability.

Future work should distinguish at least three graphs: a biological pathway graph, a measurement-process graph derived from shared analytical steps, and a data-derived residual-dependence graph estimated only within training folds. A multi-graph extension of PathEQA could learn nonnegative graph weights while retaining a graph-free component, thereby allowing the data to attenuate an unhelpful prior. Additional EQA rounds would support genuine next-round forecasting, calibrated uncertainty intervals, and prospective testing of whether graph-guided warnings improve participant investigations and subsequent performance.

## 7 Conclusion

PathEQA is a feature-graph-guided random-forest framework for multianalyte quality assessment in which a user-supplied graph can influence representation, randomized candidate search, and split optimization. Catecholamine EQA was used as one worked example rather than as a restriction of the model. The method improved prediction and graph coherence in graph-aligned small-sample simulations but could be harmed by graph misspecification. In the six-analyte case study, the biochemical pathway graph changed tree structure and yielded interpretable analyte combinations and quality phenotypes, yet did not consistently outperform simpler baselines. DA-NE was the strongest pair, DA-NE-E best estimated multianalyte burden, and DA-NE-NMN best screened for any failure. The main methodological conclusion is therefore conditional: graph-guided forests can be useful when the supplied feature graph is relevant to the prediction target, and that relevance should be tested with ablations and falsification controls rather than presumed from domain plausibility.

## Data Availability

All data produced in the present study are available upon reasonable request to the authors

## Data and code availability

The participant-level EQA dataset and the custom PathEQA implementation are not publicly released in this priority-establishing preprint version. Aggregate results and evaluation summaries are reported in the manuscript. Information about access to underlying EQA records may be requested from the corresponding author and considered subject to applicable data-governance requirements. The exact algorithmic specification, pseudocode, tuning constants, and reference implementation are reserved for a later release following intellectual-property review.

